# Impact of Cardiac Position, Morphology and Operative Technique on Long-Term Fontan Outcomes in Heterotaxy

**DOI:** 10.1101/2024.06.03.24308406

**Authors:** Sachiko Kadowaki, Zeynep uçar, Chun-Po Steve Fan, Yasmin Zahiri, Kok Hooi Yap, Teerapong Tocharoenchok, Weidan Chen, Anne I. Dipchand, Osami Honjo, David J. Barron

## Abstract

**Objectives:** Heterotaxy is characterized by complex venous connections and cardiac position requiring individualized strategies for Fontan. This study aimed to assess 25-year outcomes of heterotaxy patients undergone Fontan operation, with particular focus on morphological features and surgical techniques.

**Methods:** 82 consecutive heterotaxy patients who underwent Fontan operation from 1985 to 2021 were compared to 150 with tricuspid atresia (TA) and 144 with hypoplastic left heart syndrome (HLHS). Kaplan-Meier method and Cox proportional hazard model were used for transplant-free survival and predictor analysis.

**Results:** Right Atrial Isomerism (RAI) was present in 45 cases and Left Atrial Isomerism (LAI) in 37. The 20-year transplant-free survival was comparable between the groups (RAI vs. LAI, 76% [95% confidence interval, 57-87%] vs. 68% [47-82%], p=0.22). Techniques were extracardiac in 66%, intra-atrial conduit in 9%, lateral tunnel in 18%. More intervention on pulmonary veins occurred in the RAI group. Cardiac position and apicocaval juxtaposition did not influence outcome. The IVC-contralateral PA Fontan was associated with 100% survival, while the IVC-ipsilateral PA Fontan at the side of the cardiac apex showed 67% [34-87%] survival at 20 years. Moderate or severe ventricular dysfunction at 20 years was found in 15% (6-39%) and 0% of RAI and LAI patients among hospital survivors, respectively (p=0.09). In-hospital mortality was higher in heterotaxy (9.8% [5-19%]) compared to TA (1.3% [0.3-5.3%], p<.01) and HLHS (2.8% [1.1-7.3%], p=0.02). There was no in-hospital death after 2000 in any of groups. The 20-year transplant-free survival in heterotaxy (72% [59-82%]) was worse than that in TA (95% [89-97%], p<0.001) but was not significantly different from that in HLHS (80% [69-87%], p=0.11). Reintervention rate at 20 years was significantly higher in heterotaxy (18% [11-29%]) vs. HLHS (8% [4-16%]) (p=0.01). Ventricular dysfunction was a predictor for death (univariate p<0.001).

**Conclusion:** Various routing techniques can be successfully applied to overcome the anatomical challenges of heterotaxy. Although higher in-hospital mortality and reintervention rate were found in heterotaxy in the early era, overall survival post Fontan was similar to that in HLHS. RAI had comparable survival to LAI with greater proportion requiring pulmonary vein intervention at time of Fontan.

## Introduction

Heterotaxy is a challenging surgical subtype in congenital heart disease, characterized by abnormal systemic and pulmonary venous (PV) connections, unusual cardiac position, such as apicocaval juxtaposition (ACJ), and high incidence of common atrioventricular (AV) valve [1]. More than 75% of heterotaxy cases have a functional single ventricle, and so are destined for staged single ventricle palliation and subsequent Fontan operation [2,3]. The associated extracardiac anomalies add further challenges to the already suboptimal single ventricle circulation, including hepatic disease and ciliary dyskinesia [4,5]. Consequently, Fontan palliation in heterotaxy, right atrial isomerism (RAI) in particular, is renowned for higher mortality and morbidity [6-12].

Reports of Fontan outcomes in heterotaxy have been hampered by small case numbers, selection biases, variations in follow-up duration, and a lack of comparable non heterotaxy Fontan counterpart [13-17]. These limitations are summarized in a recent meta-analysis that identified 21 studies, but the median number of patients per study was only 24 with median follow-up of 4 ½ years [18]. The meta-analysis found a significantly higher early mortality (14%) with a 10-year survival of 74%. A notable finding was that early mortality remains high even among the recent studies, whereas survival has been improved in other anatomic subgroups undergoing Fontan operation [18].

There are few large single centre studies in which consistent techniques for the Fontan procedure have been used [17,19]. However, the role of the different techniques of Fontan modification in heterotaxy have never been studied in detail with respect to their influence on long-term outcomes. This study aimed to analyze the impact of cardiac structures and individualized operative approaches, including a low threshold to intervene on PV stenosis in the setting of RAI, at the time of Fontan operation. To assess these outcomes in context, the heterotaxy group were compared with two contemporaneous cohorts, the ‘ideal’ Fontan (Tricuspid Atresia cohort, TA), and a higher risk Fontan group (Hypoplastic Left Heart Syndrome cohort, HLHS) [20-22].

## Patients and Methods

The study protocol was approved by the Research Ethics Board, the Hospital for Sick Children, Toronto, Ontario, Canada (ID 1000071680). The cardiovascular surgery database at the Hospital for Sick Children was reviewed for all patients with heterotaxy syndrome, TA, and HLHS undergone a Fontan operation. Among the 551 patients who underwent Fontan operation between January 1985 to September 2021: 82 with heterotaxy (15%), 150 with TA (27%), and 144 with HLHS (26%) were reviewed. Heterotaxy was diagnosed in preoperative imaging modalities, including echocardiography, magnetic resonance imaging and computed tomography, and by intraoperative findings.

Data collected included demographics, underlying anatomy, operative details, and long-term follow-up data. The analysis included the subset of patients with hetarotaxy, subdivided into those with RAI (n=45) and those with left atrial isomerism (LAI) (n=37). Reoperation was defined as all cardiac surgical interventions except for permanent pacemaker implantation, generator change, and heart transplantation (HTx). The term “the IVC” in the manuscript is defined as the IVC or hepatic veins (HVs) with interrupted IVC draining into the atrium. ACJ was a morphologic feature of the cardiac apex pointing toward the ipsilateral side of the IVC [23]. The IVC-ipsilateral pulmonary artery (PA) Fontan indicates a Fontan route placed between the IVC and ipsilateral PA at the side of the cardiac apex or a Fontan route running between the IVC draining into the mid-atrium to the PA behind the ventricular mass, as opposed to the standard Fontan, in which a Fontan route is placed in the opposite side of the cardiac apex between the IVC and ipsilateral PA. The IVC-contralateral PA Fontan represents an extracardiac Fontan conduit passing over the vertebra to run to the contralateral PA. Although it has been routine to create a fenestration since early 2000’s, the decision to create a fenestration in heterotaxy was individualized based on multiple factors, including physiology, anatomical limitations, and surgical judgement. A fenestration was typically not created in patients with arteriovenous malformation (AVM) or who previously underwent Kawashima procedure. Early death or reoperation are defined as death or surgical intervention within one-month post-Fontan, while late death represents death later than one-month post-Fontan.

## Data analysis

Clinical characteristics at the index surgery were summarized using descriptive statistics. Continuous variables were summarized using median, the first and third quartile, categorical variables were summarized by frequencies. The freedom from death or HTx was characterized using the Kaplan-Meier survival method, and between-group differences in HTx-free survival were evaluated using log rank tests. The length of stay (LOS), reoperation and the incidence of moderate or greater systolic dysfunction and AV valve regurgitation (AVVR) were characterized using Fine and Gray’s subdistribution method, in which death or HTx was considered a competing risk, in terms of cumulative incidence function (CIF). Between-group differences in CIF were evaluated using Gray’s tests.

The associations of heterotaxy with the HTx-free survival was assessed and quantified using a Cox proportional hazard model in terms of hazard ratios (HRs). The association of heterotaxy with the LOS, reoperation and the incidence of moderate or greater systolic ventricular dysfunction and AVVR using the subdistribution regression method was quantified in terms of subdistribution HRs. We conducted each analysis with and without covariate adjustment. The covariates included the year of the Fontan, age and sex at the Fontan, the presence of fenestration, and other concomitant AV valve repair. In all regression analyses, the year of the Fontan and age at the Fontan, were modeled using natural cubic splines to account for their potential nonlinear associations with the outcomes; hence, the corresponding results were shown graphically. The corresponding 95% confidence interval (CI) and p-values was evaluated using Wald’s statistics. Finally, we conducted an exploratory analysis to assess clinical risk factors specified *a priori* on HTx-free survival using a Cox proportional hazard model. The risk factors under consideration included ventricular morphology, laterality of the apex of the heart, laterality of Fontan pathway, the IVC on the same side as the apex, repair of pulmonary venous obstruction, AV valve repair and Fontan route on the same side as the apex. Missing values were imputed using multivariable imputation with a chained equation. The imputed results were pooled using Rubin’s method. Statistical analyses assumed a significance level of 5% and were conducted in R v4.0.3.

## Results

The median (Q1-Q3) follow-up was 11 (3.9-20), 12 (5.3-24), and 13 (4.3-18) years for heterotaxy, TA, and HLHS, respectively. Clinical characteristics of these cohorts are described in Table S1.

### Anatomical Features and Fontan Techniques in Heterotaxy

Baseline characteristics, anatomical features, Fontan techniques in the RAI and LAI are shown in Table 1. Representative figures of Fontan pathway in each technique: standard extracardiac, intra-atrial conduit, IVC-ipsilateral PA, and IVC-contralateral PA Fontan, are shown in Figure 1. Applied surgical approaches according to locations of the IVC to the cardiac apex are summarized in Figure S1. Atrioventricular septal defect was present in most patients (76%) and in similar rate between RAI and LAI. The dominant ventricle was RV in 50% of RAI and in 70% of LAI. Dextrocardia was present in 29% of RAI and in 46% of LAI. ACJ was present in 22% of the whole cohort.

**Figure 1.**
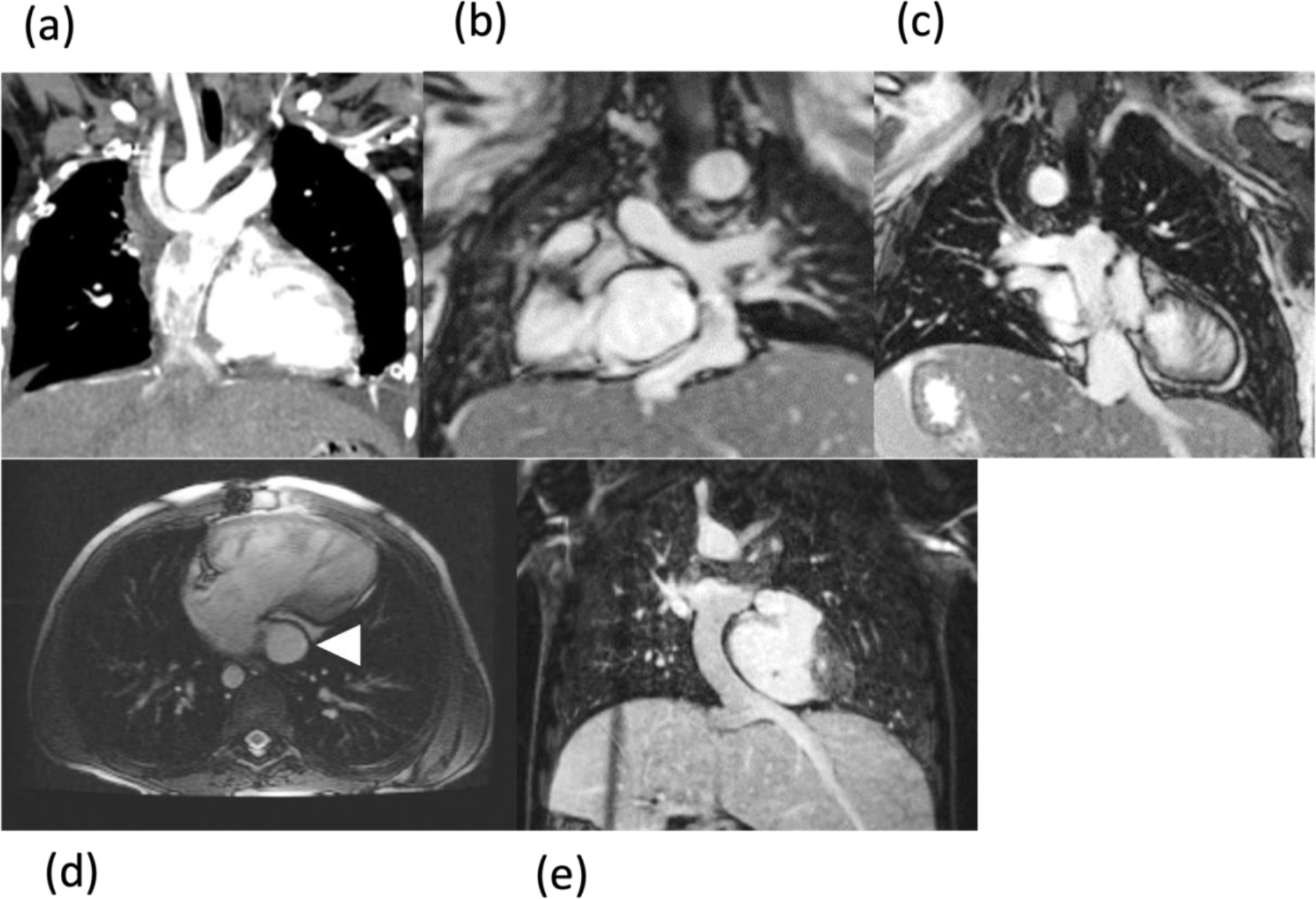
Fontan modifications. (a) right-sided extracardiac Fontan, (b) left-sided extracardiac Fontan, (c) intra-atrial conduit Fontan, (d) the IVC-ipsilateral PA Fontan, in which the arrow head indicates the conduit running behind the ventricular mass and on the same side as the cardiac apex, (e) the IVC-contralateral PA Fontan. IVC, inferior vena cava; PA, pulmonary artery.

**Table 1.**
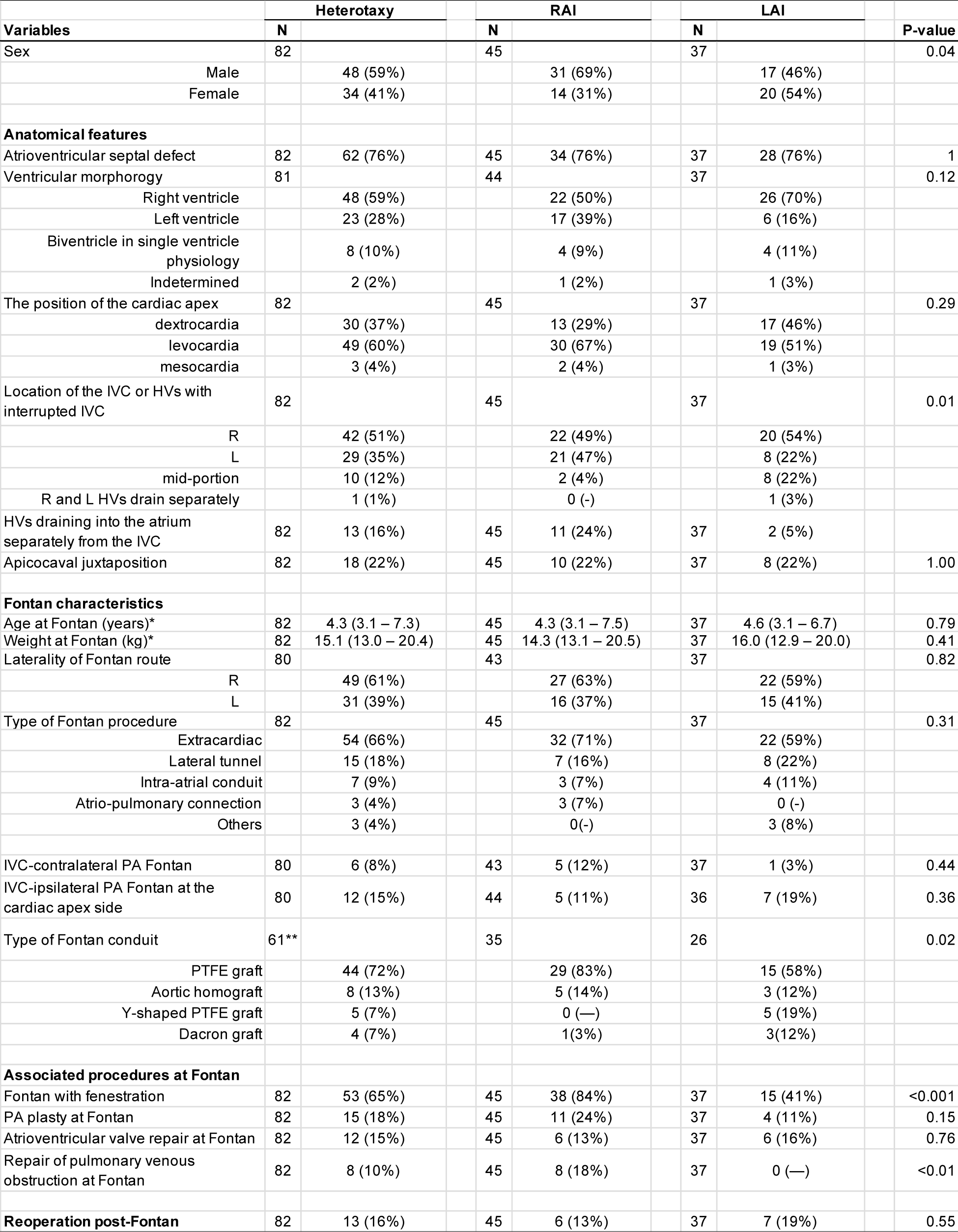
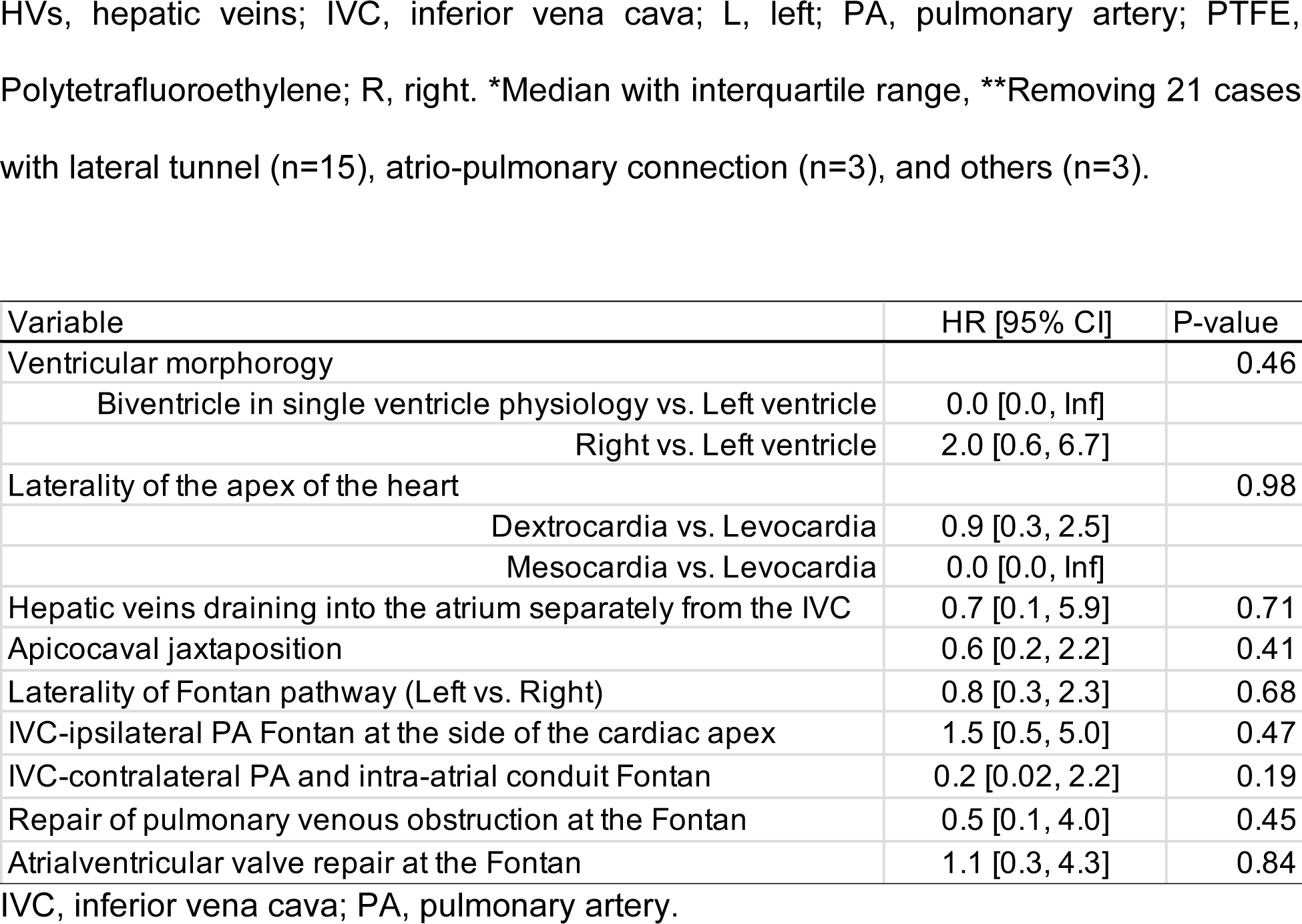
Demographic and operative variables in RAI and LAI.

The IVC was interrupted in 84% of LAI and in 2% of RAI. All patients with interrupted IVC underwent Kawashima procedure. There was no difference in age at Fontan completion between the patients who previously underwent Kawashima procedure and the patients who did not (age in years at the Fontan completion with vs. without previous Kawashima, 4.7 [3.2-7.3] vs. 4.2 [3.1-6.8], years old, p=0.82). The IVC (or HVs in the setting of interrupted IVC) drained to the right-sided atrium in approximately 50%, to the left-sided atrium in 35%, and to the midline in 12%.

An extracardiac Fontan was the commonest technique, used in 66% of the cases (71% RAI and 59% LAI) with a polytetrafluoroethylene (PTFE) graft (GORE-TEX Stretch Vascular Graft, GORE, Flagstaff, AZ) with the size ranging from 16 to 22mm. A Y-graft was used in 5 cases (all LAI) who had unilateral or bilateral AVM to ensure IVC flow to both lungs. The second commonest Fontan type was a lateral tunnel, used in 18% of cases, which were equally distributed in both RAI and LAI. An intra-atrial conduit Fontan was used in seven cases (9%) (no difference in RAI vs. LAI). Of which two woven double velour graft (20 and 24mm, respectively, Hemashield, GETINGE, Göteborg, Sweden) was used and a PTFE graft (16 to 18 mm) was used in 5 patients in order to avoid pulmonary venous compression typically in cases of midline IVC or ACJ (see Fig 1c). Three patients in the very early cohort had an atrio-pulmonary connection.

In terms of laterality of the Fontan route, 37% of RAI and 41% of LAI patients had a left sided conduit. This was strongly correlated with the incidence of dextrocardia (37%) and with the IVC (or HVs in the case of interrupted IVC) draining into the left-sided atrium (35%). There were six (8%) and 12 patients (15%) undergone IVC-contralateral PA Fontan and IVC-ipsilateral PA Fontan, respectively.

### Concomitant Procedures

Fenestration was performed in 65% of cases and was commoner in RAI (84%) than LAI (41%) (p<0.001). PV repair was performed in 18% of RAI patients and 0% of LAI (p<0.001) with zero recurrence of PV obstruction postoperatively. PA plasty and AV valve repair were performed for 18% and 15% of the patients, respectively, without significant difference between RAI and LAI.

### Control Groups: TA and HLHS

Clinical characteristics of the comparison cohorts with TA or HLHS undergoing Fontan are described, in Table S1. In comparison to patients with TA and HLHS, weight and age at the Fontan for patients with heterotaxy were larger and older. Fenestration creation was more common in the HLHS cohort than heterotaxy.

### Early outcomes

The competing risk analysis showed that hospital (4-week) mortality (95% CI) was significantly higher in heterotaxy (9.8% [5.1-19%]) than TA (1.3% [0.3-5.3%], p<0.01) or HLHS (2.8% [1.1-7.3%], p=0.02) patients, while there was no significant difference between RAI and LAI (11% [4.3-27%] vs. 8.9% [3.5-23%], p=0.72) (Figure 2.1). The estimated proportion (95% CI) of patients discharged from hospitalization alive within 4 weeks was significantly lower in heterotaxy (79%, [71-89%]) than TA (91%, [86-95%], p=0.008) or HLHS (90%, [86-95%], p=0.003); there was no significant difference between RAI and LAI (78% [66-93%]; 80% [69-93%]; p=0.77) (Figure S2).

**Figure 2.**
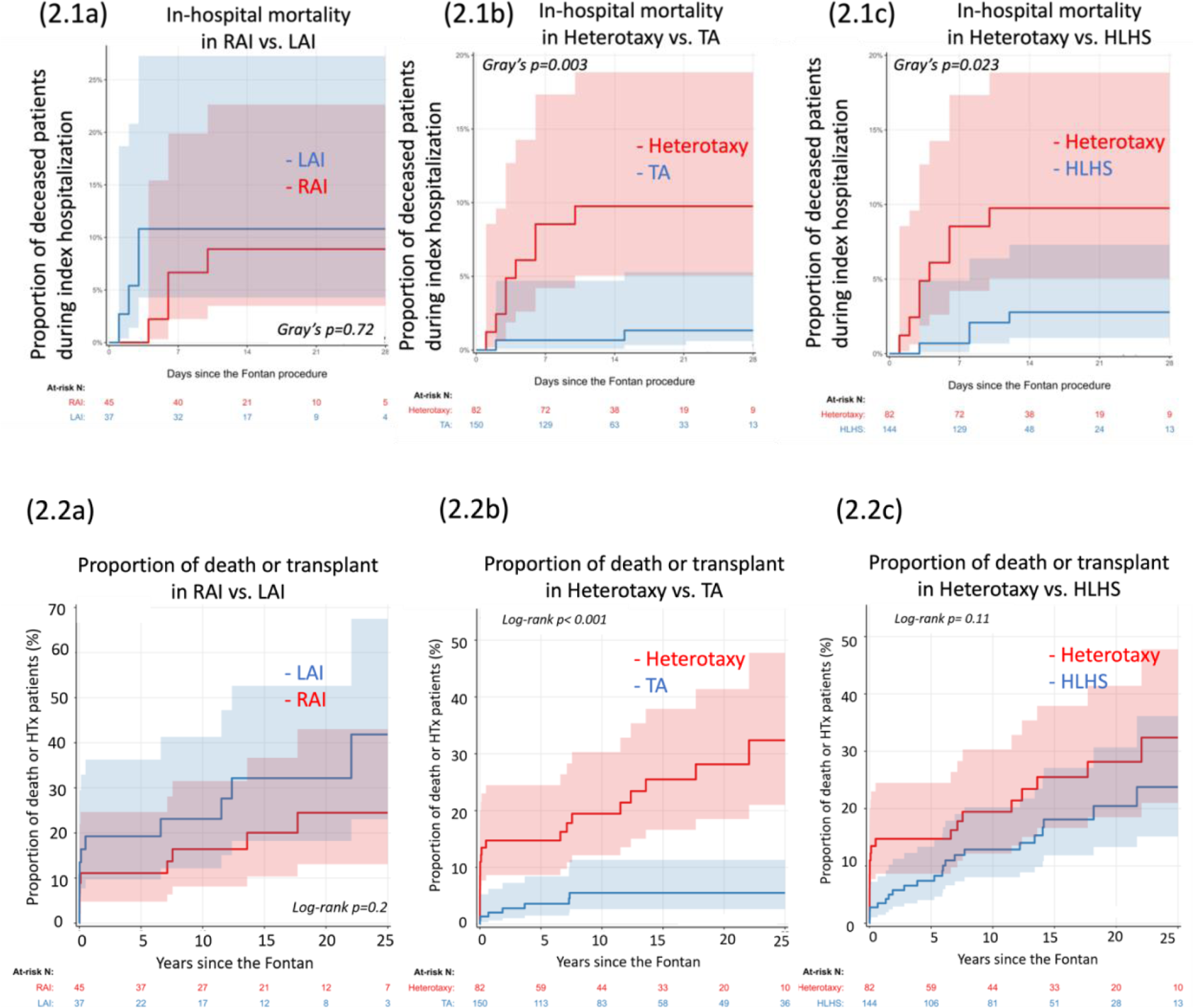
The proportion of deceased or transplanted patients after Fontan operation. The follow-up period was during hospitalization post-Fontan (2.1) and over the period post-Fontan (2.2). (a) RAI vs. LAI, (b) Heterotaxy vs. TA, and (c) Heterotaxy vs. HLHS.

Moreover, the results of subdistribution regression analysis showed that heterotaxy was significantly associated with a reduced likelihood of being discharged alive (thus more likely to die during the index hospitalization) (Table 3) compared to TA or HLHS. It was estimated that heterotaxy was associated with an approximate 40% reduction in the likelihood of a live discharge compared to TA (HR [95% CI] = 0.64 [0.47, 0.89], p = 0.007) or compared to HLHS (0.75 [0.54, 1.03], p = 0.08). It is also noteworthy that significant improvement in terms of the incidence of live discharges year over year in all groups (RAI and LAI, heterotaxy and TA, and heterotaxy and HLHS) (Figure 3) and that the covariate-adjusted effect of Fontan year on HTx-free survival was not statistically significant (Figure S3). Modes of early death in heterotaxy are summarized in Figure S4. Notably, three out of 11 early deaths occurred in the IVC-ipsilateral PA group, of which two occurred during the index hospitalization due to failed Fontan circulation, and the other patient died at home nine days post-Fontan.

**Figure 3.**
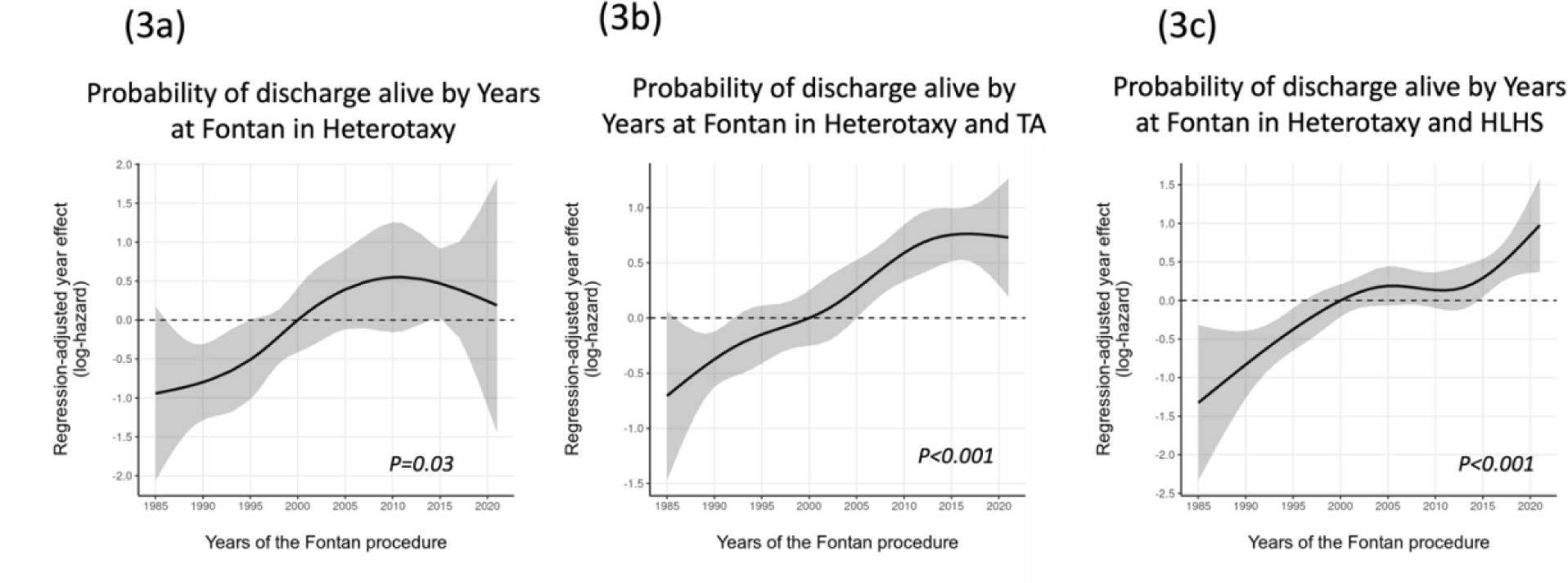
Regression-adjusted year effect for discharge alive. In the combined cohorts of RAI and LAI (a), Heterotaxy and TA (b), and Heterotaxy and HLHS (c).

Early reoperation was required for six (7.3%) heterotaxy, two (1.3%) TA, and one (0.7%) HLHS patient (Table S1, Figure 4). Among those with early reoperation, Fontan takedown was required in two heterotaxy, one TA, and one HLHS patient during the index admission with three in-hospital deaths except for one TA post-Fontan takedown.

**Figure 4.**
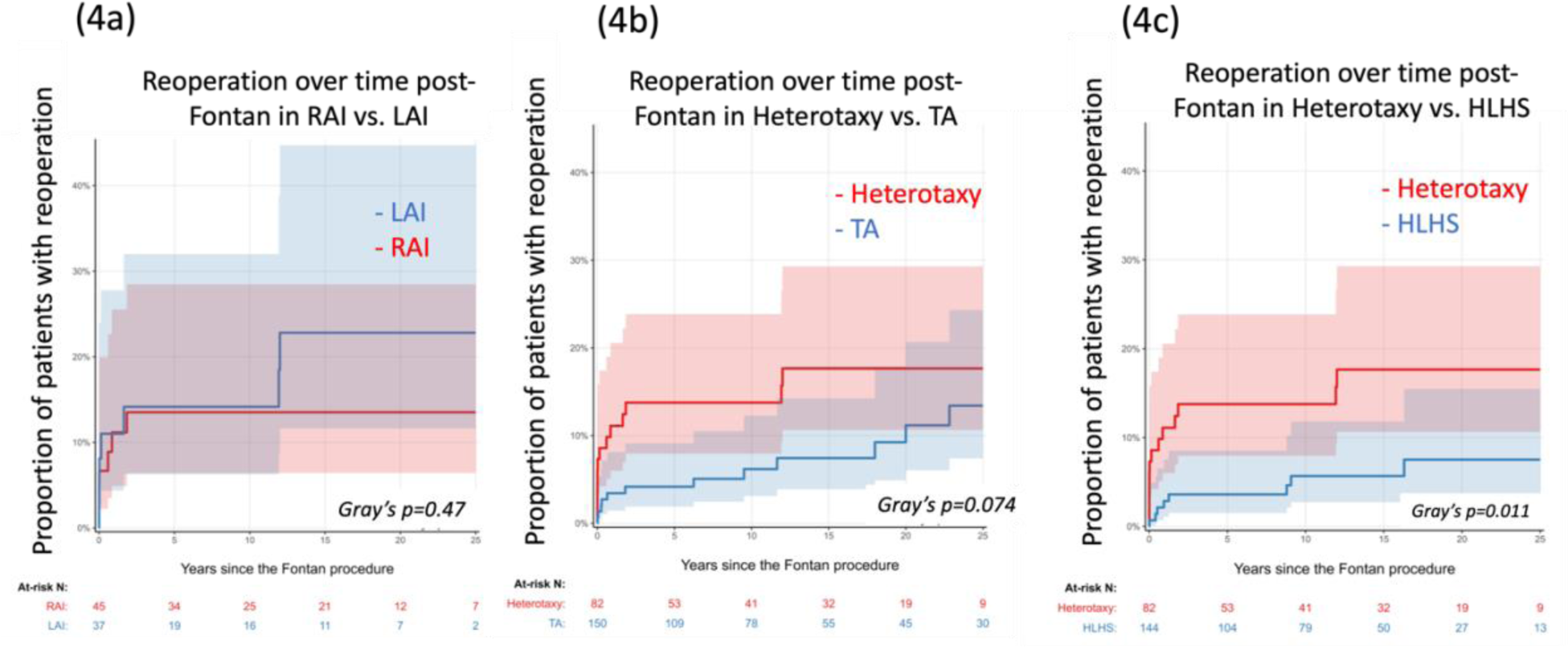
Reoperation rate over time following Fontan operation. (a) RAI vs. LAI, (b) heterotaxy vs. TA, and (c) heterotaxy vs. HLHS.

### Late outcomes

There were in total 20 heterotaxy, 8 TA and 22 HLHS patients who subsequent died or received HTx during the follow-up. The proportion of deceased or transplanted patients in each group is shown in Figure 2.2 and modes of late death or HTx are shown in Figure S4. There was no significant difference in the 20-year HTx-free survival between RAI and LAI populations. The HTx-free survival (95% CI) at 20 years after Fontan was significantly better in TA (95%, [89-97%]) than that in heterotexy (72%, [59-82%], p<0.001) and not significantly different between heterotaxy and HLHS (80%, [69-87%], p=0.11). The findings remained consistent among hospital survivors, in which the 20-year HTx-free survival remained significantly lower in heterotaxy (82% [67-90%]) than that in TA (96% [90-98%], p<0.01) and not significantly different from that in HLHS (82% [71-89%], p=0.94). The multivariable Cox proportion hazard regression showed that the risk of death or HTx was significantly higher in the heterotaxy than the TA patients (adjusted HR [95% CI] = 4.2 [1.5, 12], p = 0.007) and not significantly different from the HLHS patients (adjusted HR [95% CI] = 1.2 [0.5, 2.7], p = 0.63) (Table S2).

There were in total 13 heterotaxy, 13 TA and 8 HLHS patients who received reoperations during the follow up. The estimated proportion (95% CI) of reoperation at 20 years after Fontan in the heterotaxy group (18%, [11-29%]) was not significantly different from that in TA (11%, [6.0-21%], p=0.07) but was significantly higher than that in HLHS (7.5% [3.7-16%], p=0.01) (Figure 4). There were no significant differences in the incidence of reoperation between RAI and LAI. Moreover, late reoperation was required for seven (9%), eleven (7%), and seven patients (5%) in heterotaxy, TA, and HLHS, respectively (Table S1). One of two Fontan revisions in heterotaxy was attributed to PV obstruction caused by the intra-atrial Fontan conduit, for which mobilization of the conduit and repair of the PV obstruction were performed. Another Fontan revision was thrombectomy from the Fontan conduit. Retrieval of a fenestration closure device was needed in one patient.

### Ventricular Dysfunction and AVVR in Heterotaxy

The impact of moderate or greater systolic ventricular dysfunction and moderate or greater AVVR among the hospital survivors in the heterotaxy group are shown in Figure 5. None of the LAI patient developed moderate or greater ventricular dysfunction, while the estimated proportions of RAI patients who developed moderate or greater ventricular dysfunction were 2% (0.3-15%), 6% (2-24%), 15% (6-39%) at 10-, 15-, and 20-years post-Fontan. The proportions of patients with moderate or greater AVVR were 17% (9-34%) at 10 and 15 years and 42% (27-65%) at 20 years in RAI, while those in LAI were 6% (2-25%) at 10 years and 16% (6-40%) at 15 and 20 years. There was no statistical significance in the incidence of ventricular dysfunction (p=0.09) and AVVR (p=0.10) between RAI and LAI.

**Figure 5.**
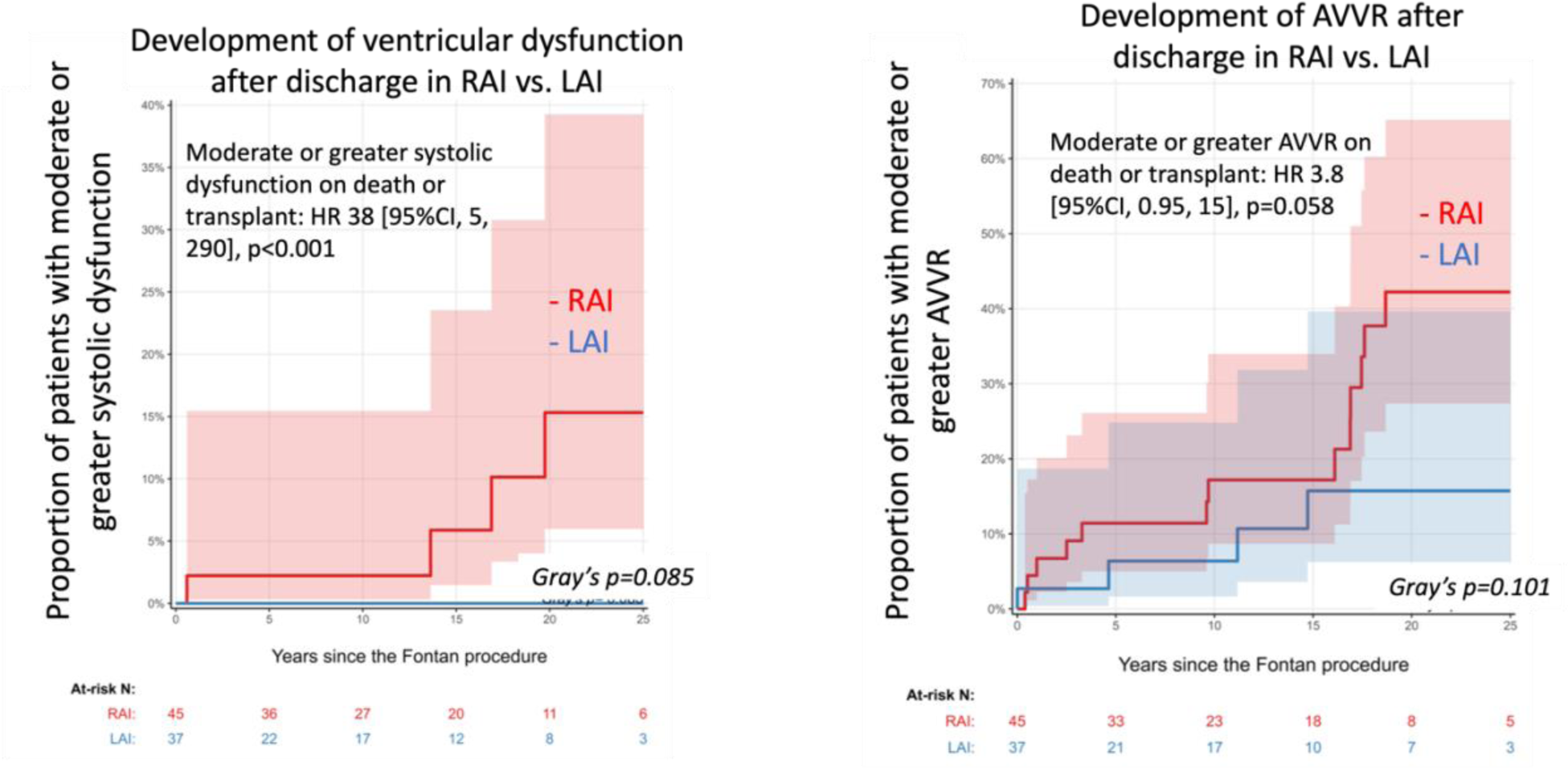
Development of systolic dysfunction and atrioventricular valve regurgitation (AVVR) in heterotaxy patients who survived to hospital discharge post-Fontan. (a) moderate or greater systolic dysfunction, (b) moderate or greater AVVR. The univariate analysis on death or transplant by development of moderate or greater ventricular dysfunction and AVVR after discharge are described in each figure. AVVR, atrioventricular valve regurgitation; CI, confidence interval; HR, hazard ratio.

### Risk Analysis for HTx-free Survival

The univariate analysis showed no anatomical and Fontan characteristics associated with HTx-free survival in heterotaxy (Table 2). The technique of routing the extracardiac conduit to the contralateral side of the cardiac mass (n=6) was associated with no early or late mortality and no pathway obstruction, although there was no statistical significance compared to other routing techniques. By contrast, there were 4 deaths out of 12 patients with IVC-ipsilateral PA Fontan over the period, showing HTx-free survival at 67% [34-87%] at 20 years.

## Discussion

This series constitutes a large single-centre experience of Fontan operation for patients with heterotaxy with median follow up of over 11 years. Despite concerns over the long-term outcomes for RAI, the study showed improved outcomes for RAI that are comparable to those of LAI. This may have been achieved through the aggressive treatment for PV stenosis at the time of Fontan operation and individualized Fontan configurations, such as the intra-atrial conduit to avoid compression on PVs. The majority of early mortality in the cohort occurred before 2000 and all three groups have universally excellent survival in the current era. Compared with HLHS, the heterotaxy group had an early hazard of death and HTx and but similar constant low late hazard, making equivalent survival between heterotaxy and HLHS among the hospital survivors.

### Individualized Fontan Techniques

The study demonstrated no significant impact of the ventricular morphology and positioning, presence of ACJ, laterality of the Fontan route, types of Fontan modifications, and requirement of concomitant PV repair or AV valve repair at Fontan on HTx-free survival in patients with heterotaxy. Although no single technique of Fontan had superior outcomes, one important message was that, in the setting of ACJ, routing an extracardiac conduit to the contralateral side to the apex showed no mortality and reoperation, while routing the conduit to the ipsilateral side to the apex was associated with some mortalities. Previous reports of Fontan for patients with ACJ showed various strategies and mixed results. A small study of seven heterotaxy patients favored ipsilateral conduit (in 77% of the cohort) with excellent survival [10] whereas the other studies favored contralateral conduit in 54% and 86% of the cohorts, respectively [12, 24]. These practice variations may stem from the efforts to mitigate the risks of PV obstruction in the IVC-ipsilateral PA Fontan and systemic venous obstruction in the IVC-contralateral PA Fontan [12,23,25,26]. To address these drawbacks of the techniques, intra-atrial conduit and intra-extracardiac Fontan techniques have gained renewed interest, even in more standard anatomy as they allow for anterior positioning of the conduit away from the PVs and can be reliably fenestrated [27-29].

### Heterotaxy vs TA and HLHS

One of the main objectives of this study is to compare outcomes of heterotaxy patients to a contemporaneous cohort of ideal and high-risk Fontan categories – TA and HLHS. We found that HTx-free survival and hospital mortality in TA were superior to those in heterotaxy, while the reoperation rate was comparable between TA and heterotaxy groups. Despite having the higher hospital mortality and reoperation rate, heterotaxy had comparable HTx-free survival to HLHS over 20 years. The era effect needs to be taken into consideration as a potential factor to alleviate early mortality in heterotaxy. Despite no statistical difference in the covariate-adjusted year effect on HTx-free survival over the 20 years (Figure S3). There has been no hospital death since 2000 in any subgroups. Therefore, future study to analyze contemporary cohort may look different than the outcomes shown in this study.

### Impact of Ventricular Dysfunction and AVVR

In univariate analysis, moderate or greater systolic dysfunction was revealed as a significant risk factor for death or HTx in patients with heterotaxy. Moderate or greater AVVR was not identified as the predictor for death or HTx in this study, which could partly be explained by the shorter follow-up period. In the longer follow-up, more patients with significant AVVR might have functional deterioration, as AVVR was identified as an independent risk factor of the long-term outcomes in previous reports [30,31]. The important facts in this study are well-maintained systolic function for the first 13 years after Fontan and steady increase in the rate of significant AVVR over time particularly among the patients with RAI. Since there is a significant negative impact of systolic dysfunction on HTx-free survival and significant AVVR can lead to ventricular functional deterioration, early intervention for AVVR in heterotaxy should be considered to prevent ventricular dysfunction and poor outcomes.

### Study limitations

There are two major limitations of this study: its retrospective nature and a small number of Fontan patients with heterotaxy, despite this being one of the largest single-centre series to date. Thus, regression analysis for short-term outcomes, including the era effect, was not conducted. Future analysis might be beneficial in identifying potential risks to early Fontan mortality and rather in describing if heterotaxy remains a high-risk group for the early post-Fontan outcome considering the era effect. In the present study, eligibility criteria for Fontan’s candidacy were not considered and patients in the heterotaxy cohort were pre-selected, excluding the ones who had not successfully undergone the Fontan completion.

## Conclusions

Heterotaxy remains a high-risk group for Fontan completion, characterized by a high early mortality compared to other Fontan substrates. The individualized Fontan routing strategy effectively prevented systemic venous and PV obstruction. Routing the conduit to the contralateral side of the ventricular mass is potentially beneficial to the patients with ACJ and related abnormalities. The potential higher risk of RAI can be mitigated by concomitant correction of any PV anomaly at time of Fontan and use of fenestration. Heterotaxy Fontan outcomes is equivalent to those of HLHS, and the late incidence of reintervention is low. Ventricular dysfunction is associated with late death and HTx.

## Data Availability

All the data can be available and need contact to the corresponding author.

## Acknowledgement

None

## Sources of Funding

No funding

## Disclosure

None

---

**Table S1.** Clinical characteristics of patients at and post-Fontan in Heterotaxy vs. TA or HLHS.

| Variables | Heterotaxy |  | TA |  | P-value* | HLHS |  | P-value** |
| --- | --- | --- | --- | --- | --- | --- | --- | --- |
|  | N |  | N |  |  | N |  |  |
| Sex | 82 |  | 150 |  | 1 | 144 |  | 0.056 |
| Male |  | 48 (59%) |  | 88 (59%) |  |  | 103 (72%) |  |
| Female |  | 34 (42%) |  | 62 (41%) |  |  | 41 (29%) |  |
| Age at Fontan (years) | 82 | 4.3 (3.1 – 7.3) | 150 | 3.1 (2.6 – 3.9) | <0.001 | 144 | 3.2 (2.5 – 3.8) | <0.001 |
| Body weight at Fontan (kg) | 82 | 15.1 (13.0 – 20.4) | 150 | 13.6 (12.0 – 15.4) | <0.001 | 144 | 13.5 (12.5 – | <0.001 |
| Fontan with fenestration | 82 | 53 (65%) | 150 | 102 (68%) | 0.66 | 144 | 124 (86%) | <0.001 |
| Reoperation post-Fontan | 82 | 13 (16%) | 150 | 13 (9%) | 0.13 | 144 | 8 (6%) | 0.02 |
| *Early reoperation* |  |  |  |  |  |  |  |  |
| Fontan takedown |  | 2 (2%) |  | 1 (0.7%) |  |  | 1 (0.7%) |  |
| Fontan revision |  | 2 (2%) |  | - |  |  | - |  |
| Pulmonary artery plasty |  | - |  | 1 (0.7%) |  |  | - |  |
| Subaortic stenosis repair |  | 1 (1%) |  | - |  |  | - |  |
| *Late reoperation* |  |  |  |  |  |  |  |  |
| Fontan takedown |  | - |  | 1 (0.7%) |  |  | 2 (1%) |  |
| Fontan revision |  | 2 (2%) |  | 1 (0.7%) |  |  | 3 (2%) |  |
| Fontan conversion |  | - |  | 6 (4%) |  |  | - |  |
| Fenestration closure device retrieval |  | 1 (1%) |  | - |  |  | - |  |
| Atrioventricular valve repair |  | 2 (2%) |  | 1 (0.7%) |  |  | 1 (0.7%) |  |
| Atrioventricular valve replacement |  | 1 (1%) |  | - |  |  | - |  |
| Pulmonary artery plasty |  | - |  | 1 (0.7%) |  |  | - |  |
| Pericardial drainage |  | 1 (1%) |  | - |  |  | - |  |
| Arterio-venous fistula creation |  | 1 (1%) |  | - |  |  | - |  |
| Bentall |  | - |  | 1 (0.7%) |  |  | 1 (0.7%) |  |

\* Heterotaxy vs. TA, \*\*Heterotaxy vs. HLHS

**Table S2.**
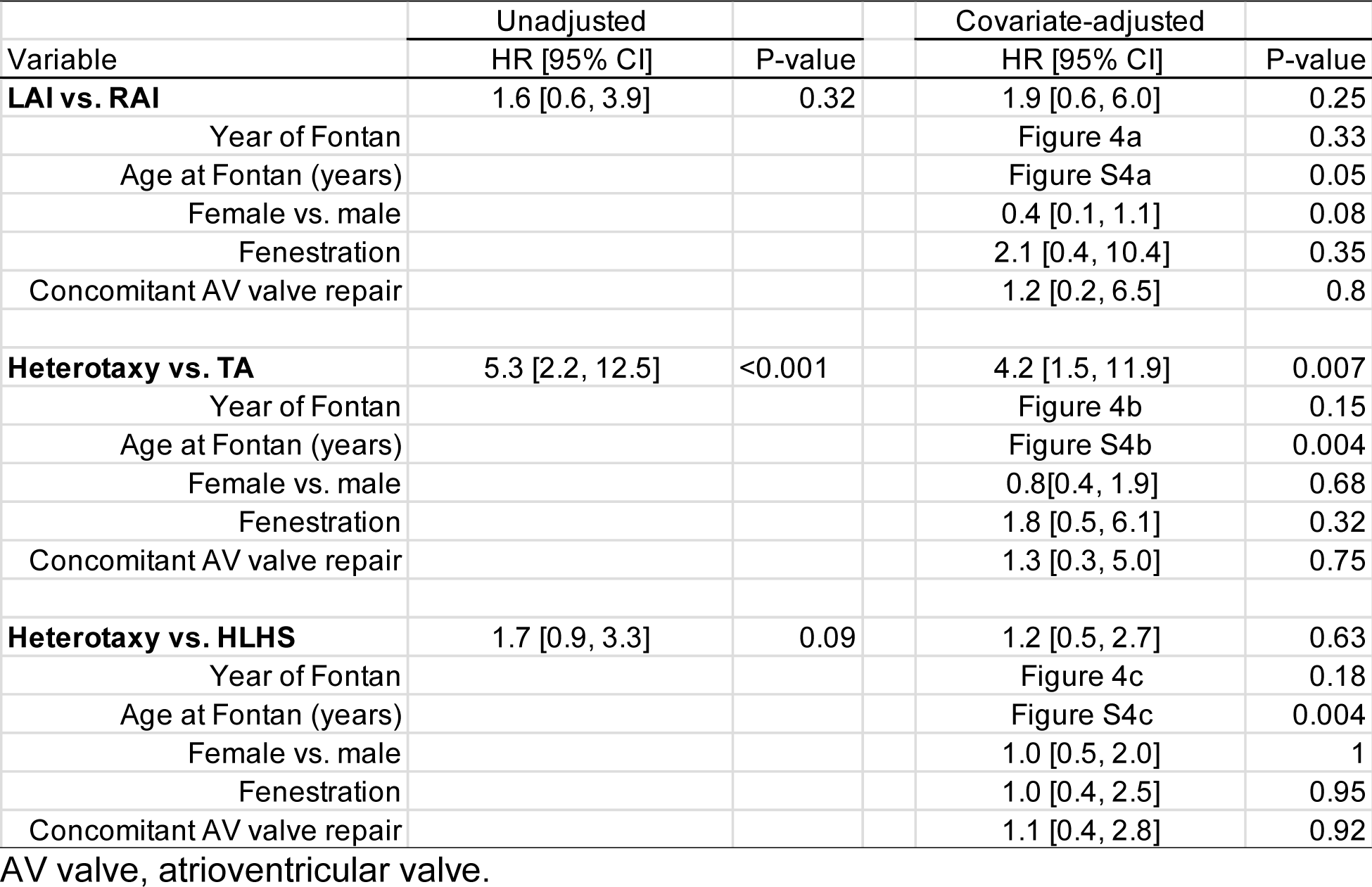
The regression results on transplant-free survival.

**Table S3.** The regression results on discharge alive.

| Variable | Unadjusted |  | Covariate-adjusted |  |
| --- | --- | --- | --- | --- |
|  | Subdistrict HR [95% CI] | P-value | Subdistrict HR [95% CI] | P-value |
| **LAI vs. RAI** | 0.9 [0.6, 1.5] | 0.72 | 0.6 [0.3, 1.0] | 0.07 |
| Year of Fontan |  |  | Figure S5.1a | 0.03 |
| Age at Fontan (years) |  |  | Figure S5.2a | 0.54 |
| Female vs. male |  |  | 2.2 [1.2, 3.8] | <0.01 |
| Fenestration |  |  | 0.5 [0.2, 1.0] | 0.06 |
| Concomitant AV valve repair |  |  | 0.6 [0.3, 1.2] | 0.15 |
| **Heterotaxy vs. TA** | 0.6 [0.5, 0.8] | 0.001 | 0.6 [0.5, 0.9] | <0.01 |
| Year of Fontan |  |  | Figure S5.1b | <0.001 |
| Age at Fontan (years) |  |  | Figure S5.2b | 0.5 |
| Female vs. male |  |  | 1.2 [0.9, 1.6] | 0.21 |
| Fenestration |  |  | 0.8 [0.6, 1.2] | 0.3 |
| Concomitant AV valve repair |  |  | 0.7[0.4, 1.4] | 0.38 |
| **Heterotaxy vs. HLHS** | 0.6 [0.5, 0.8] | <0.001 | 0.7 [0.5, 1.0] | 0.08 |
| Year of Fontan |  |  | Figure S5.1c | <0.001 |
| Age at Fontan (years) |  |  | Figure S5.2c | 0.36 |
| Female vs. male |  |  | 1.1 [0.8, 1.5] | 0.54 |
| Fenestration |  |  | 0.6 [0.4, 0.9] | <0.01 |
| Concomitant AV valve repair |  |  | 0.8 [0.6, 1.3] | 0.39 |

AV valve, atrioventricular valve.

**Figure S1.**
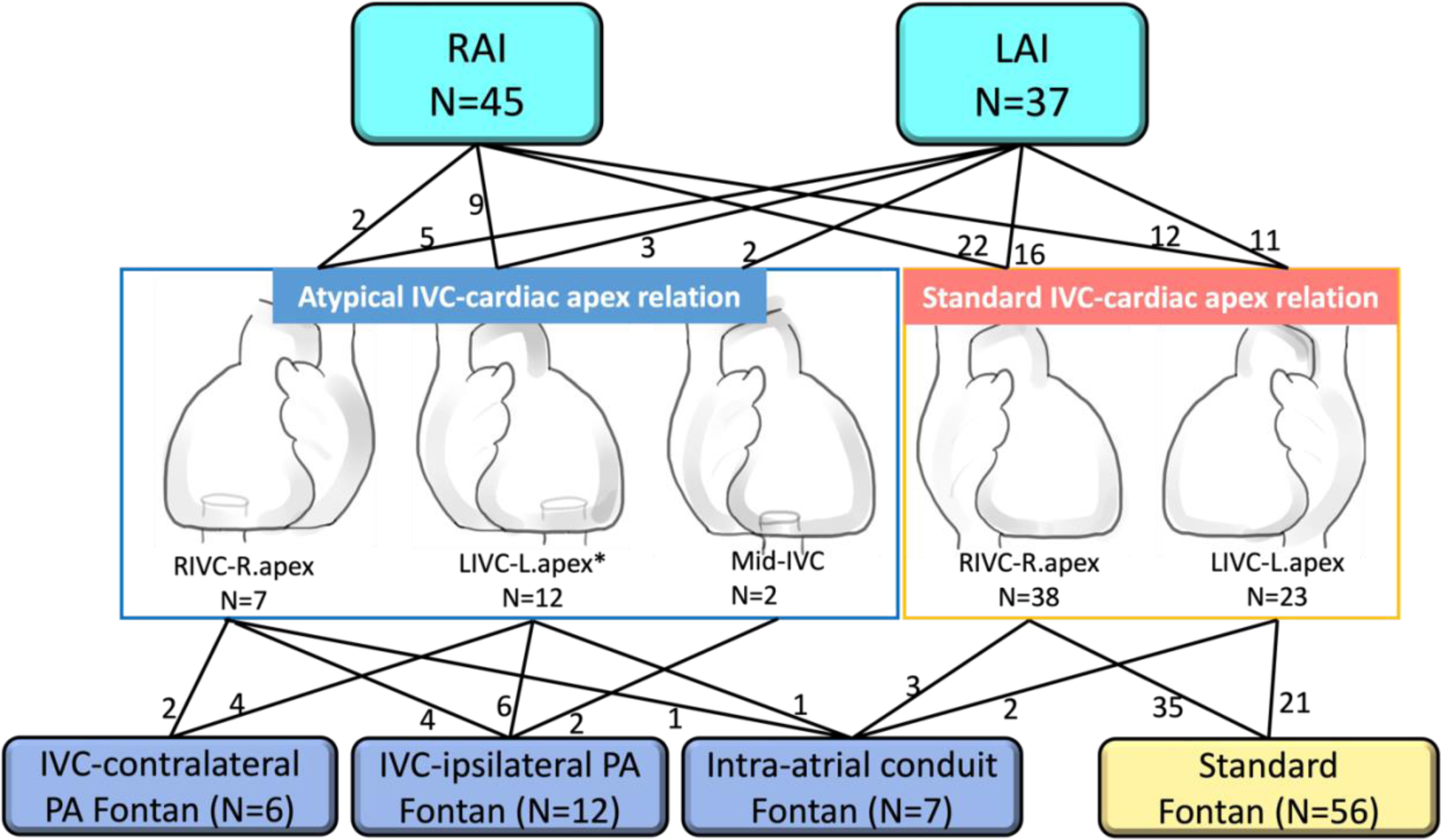
Surgical approaches using IVC-contralateral PA Fontan, IVC-ipsilateral PA Fontan, and intra-atrial conduit Fontan. *One of the patients with LIVC-L.apex relation is unknown which Fontan modification was used. HV, hepatic vein; IVC, inferior vena cava; L, left; PA, pulmonary artery; R; right.

**Figure S2.**
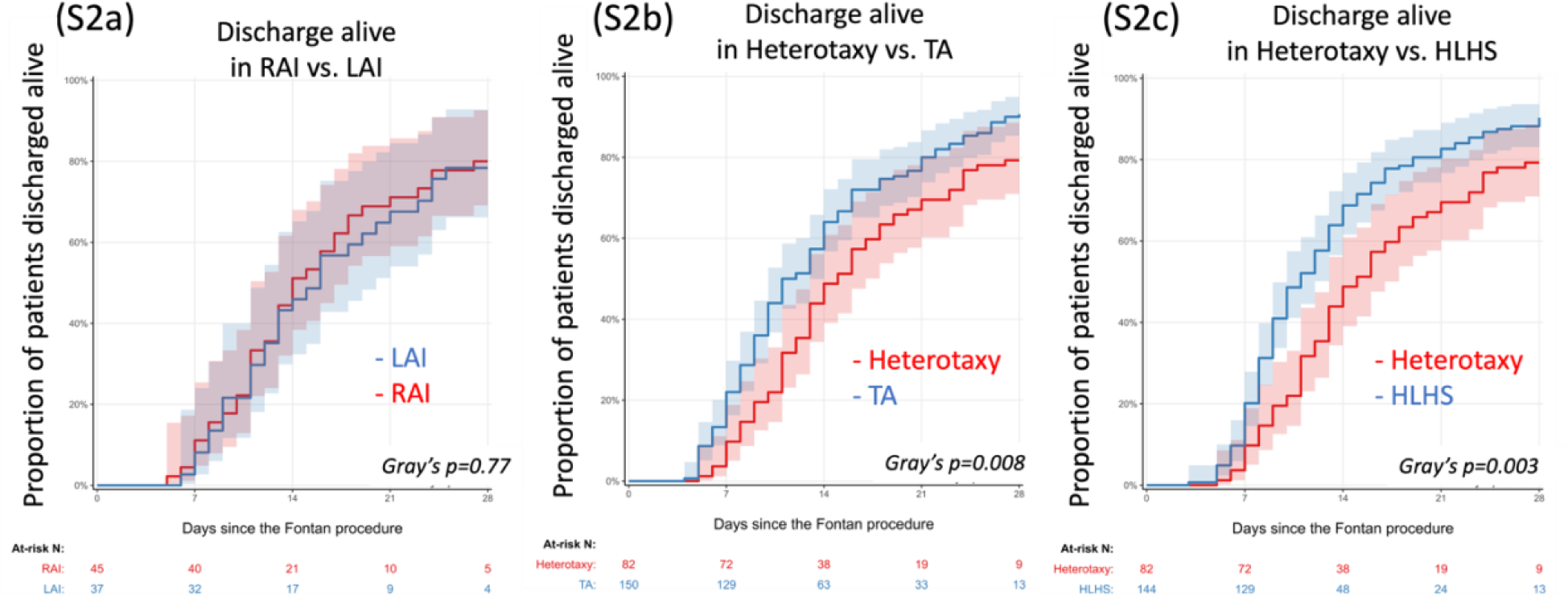
Discharge alive within a month post-Fontan. (a) RAI vs. LAI, (b) heterotaxy vs. TA, and (c) heterotaxy vs. HLHS.

**Figure S3.**
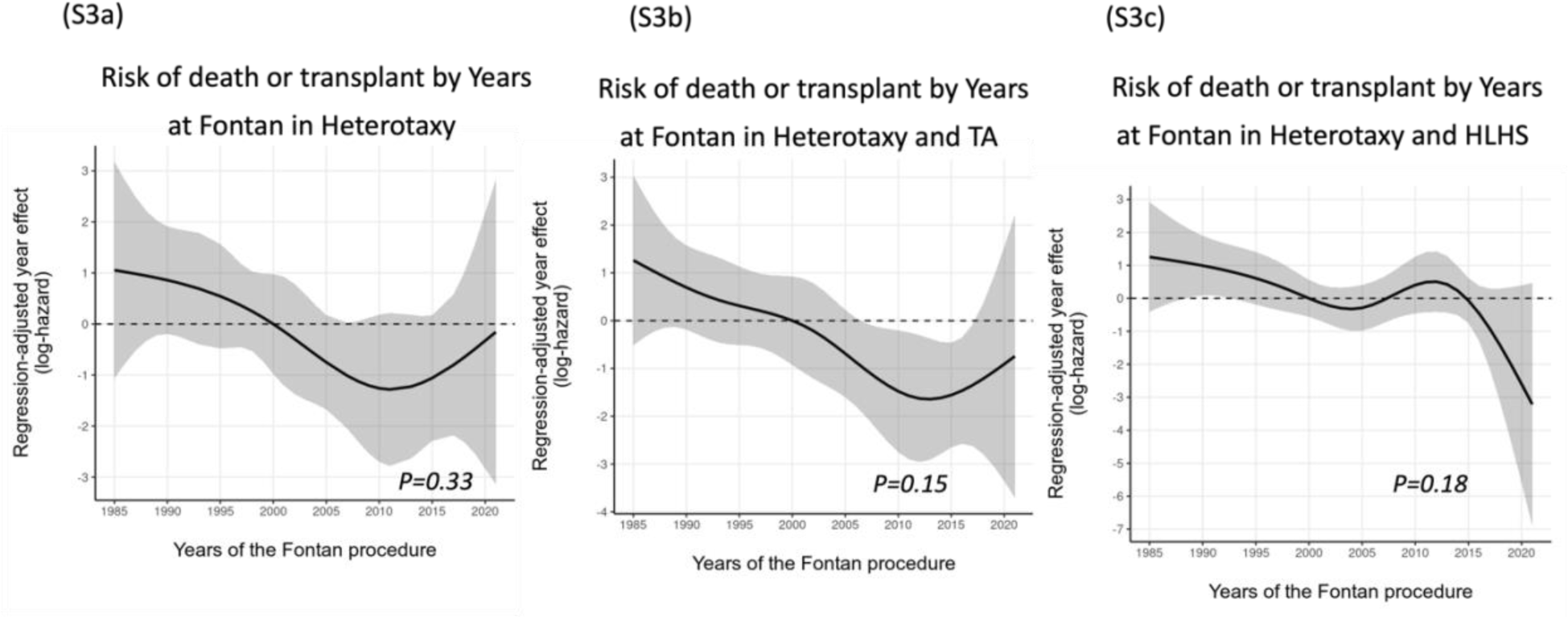
Regression-adjusted year effect for transplant-free survival. In the combined cohorts of RAI and LAI (a), Heterotaxy and TA (b), and Heterotaxy and HLHS (c).

**Figure S4.**
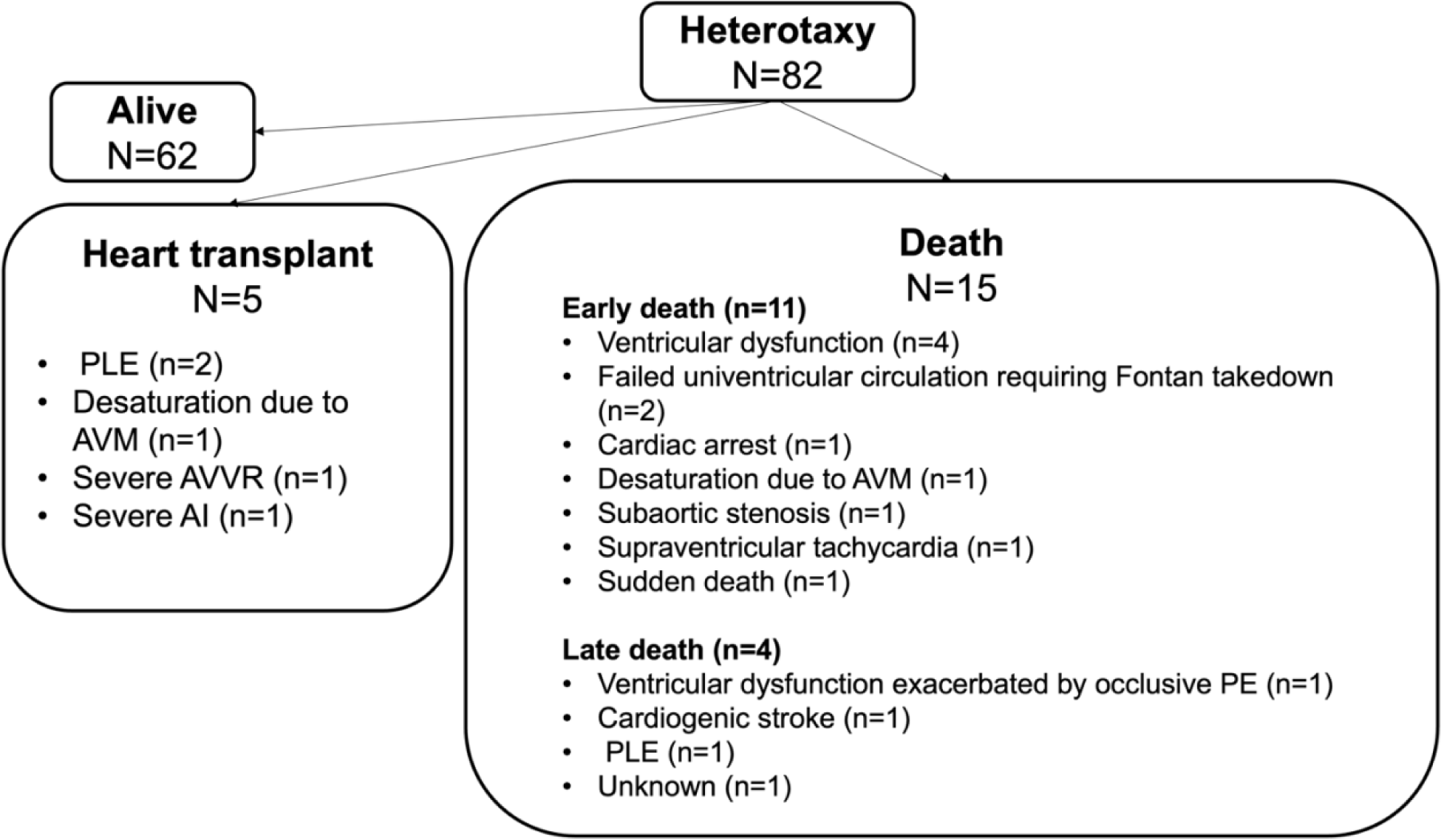
Mode of death or heart transplant post-Fontan in the heterotaxy cohort. AI, aortic insufficiency; AVM, arteriovenous malformation; AVVR, atrioventricular valve regurgitation; PE, pulmonary thromboembolism; PLE, protein losing enteropathy.

**Figure S5.**
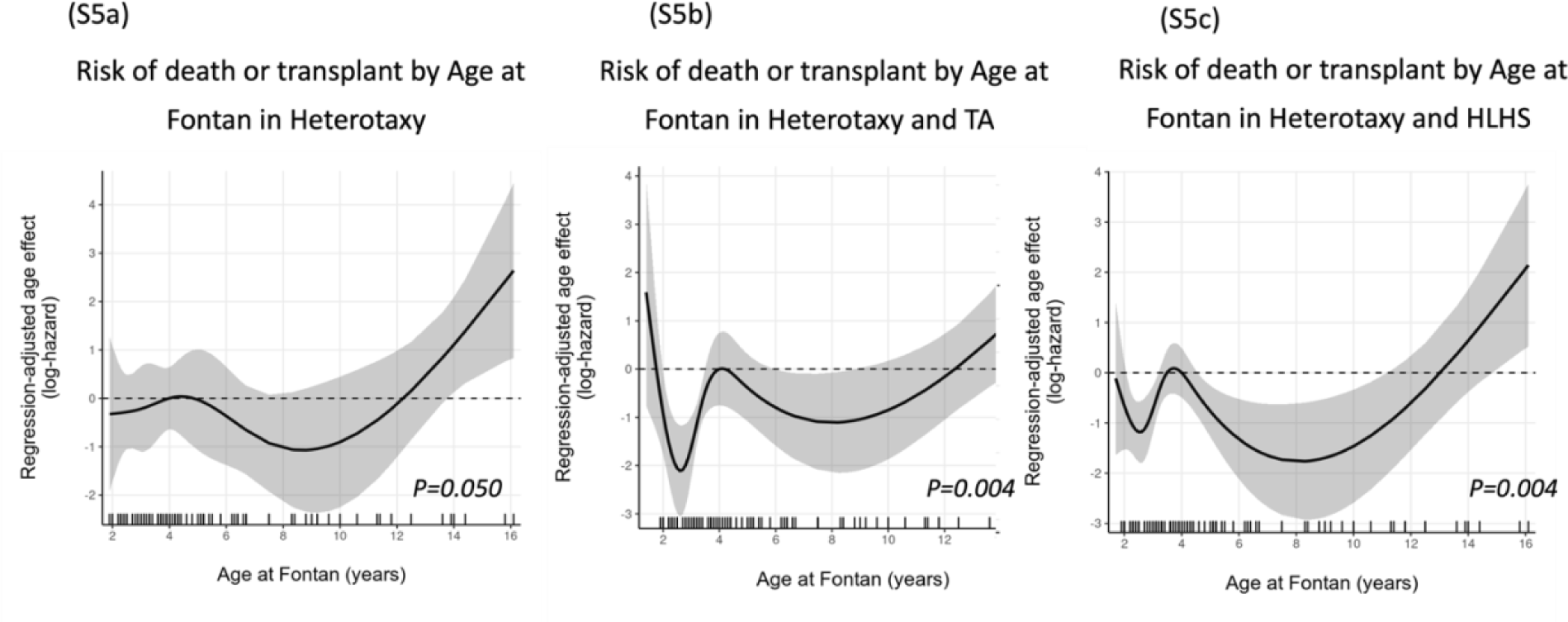
Regression-adjusted age effect for transplant-free survival. In the combined cohorts of RAI and LAI (a), Heterotaxy and TA (b), and Heterotaxy and HLHS (c).

**Figure S6.**
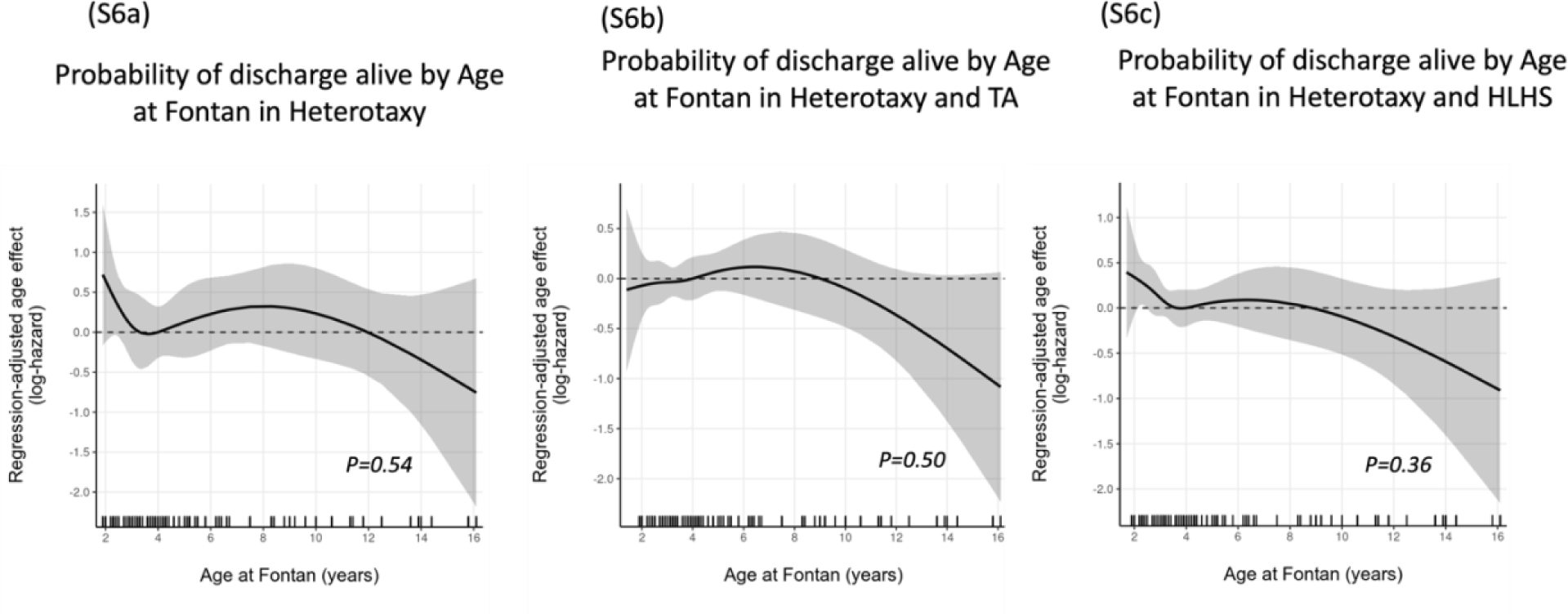
Regression-adjusted age effect for discharge alive. In the combined cohorts of RAI and LAI (a), Heterotaxy and TA (b), and Heterotaxy and HLHS (c).

## Notes

### Competing Interest Statement

The authors have declared no competing interest.

### Author Declarations

The study protocol was approved by the Research Ethics Board, the Hospital for Sick Children, Toronto, Ontario, Canada (ID 1000071680).

